# Adaptation and validation of screening measures of anxiety (GAD-7), depression (PHQ-9), and post-traumatic stress disorder (PC-PTSD-5) for use in population-based epidemiological studies in Malawi, Africa

**DOI:** 10.64898/2026.07.21.26358551

**Authors:** Rebecca Hendrine Nzawa-Soko, Robert C Stewart, Jullita Kenala-Malava, Japhet Myaba, Manson Msiska, Madalitso Makhalira, Tommy Mthepheya, Andrew Jackie Matchado, Joseph Mkandawire, Thandile Nkosi, Innocent Nyanjagha, Eric Umar, Wisdom Petros Nakanga, Louise Coombes, Nadine Seward, Angus MacBeth, Andrew Mark McIntosh, Amelia C Crampin

## Abstract

Population-based studies of common mental health conditions (anxiety, depression, post-traumatic stress disorder) require measures that are valid in the study context. We set out to validate the Generalised Anxiety Disorder-7 scale (GAD-7), Patient Health Questionnaire-9 (PHQ-9); and Primary Care PTSD Screen for DSM-5 (PC-PTSD-5) in the most widely spoken languages in Malawi (Chichewa and Chitumbuka). We undertook translation, adaptation, and piloting to produce final versions in both languages. We conducted criterion validation of the GAD-7, PHQ-9 and PC-PTSD-5 against reference diagnoses of DSM-5 generalised anxiety disorder, major/minor depressive episode, and PTSD respectively, using the Structured Clinical Interview for DSM-5 (SCID-5). A weighted sample of screened participants had SCID interview and this was adjusted for in the analysis. We recruited convenience samples of women and men from two sites: a rural Chitumbuka-speaking site where 342 were screened and 219 had SCID; and an urban Chichewa-speaking site where 458 were screened and 251 had SCID. In both languages, the measures had acceptable internal consistency (Cronbach’s alpha ≥ 0.75). Regarding convergent validity, PHQ-9 and GAD-7 were highly correlated but PC-PTSD-5 was only weakly/moderately correlated with the other measures. In Confirmatory Factor Analysis, best fit for GAD-7 and PC-PTSD-5 was a 1-factor structure, and for PHQ-9 was a 2-factor structure; there was only partial measurement invariance between the 2 language versions of each measure. Area under the ROC curve (AUC) for GAD-7 detection of generalised anxiety disorder was: Chitumbuka 0.759 (95%CI: 0.634, 0.871); Chichewa 0.868 (95%CI: 0.812, 0.915). AUC for PHQ-9 detection of major depression was: Chitumbuka 0.634 (95%CI: 0.441, 0.869); Chichewa 0.843 (95%CI: 0.721, 0.927). AUC for PHQ-9 detection of minor-or-major depression was: Chitumbuka 0.751 (95%CI: 0.619, 0.865); Chichewa 0.801 (95%CI: 0.714, 0.879). AUC for PC-PTSD-5 detection of PTSD was: Chitumbuka 0.682 (95%CI: 0.519, 0.853); Chichewa 0.741 (95%CI: 0.604, 0.859). In conclusion, GAD-7 and PHQ-9 showed good/acceptable validity, although criterion validity of PHQ-9 for major depression in Chitumbuka was poor. PC-PTSD-5 showed limitations to its validity, indicating need for further development of PTSD measures in Malawi.

## Background

Common mental health conditions including anxiety, depression and post-traumatic stress disorder (PTSD) are recognised as important global health problems [1] but there remain many gaps in the epidemiological evidence base. These gaps are particularly prominent in low- and middle-income countries (LMICs), many of which are undergoing marked demographic, health and environmental change [2]. The *Generation Malawi* and *Healthy Lives Malawi* longitudinal population-based cohort studies are addressing this evidence gap in Malawi, a low-income African nation [3].

Large population-based epidemiological studies require the use of mental health measures that are acceptable, feasible, and valid in the study context. Brief mental health screening tools, such as the Patient Health Questionnaire 9-item (PHQ-9) for depression [4] are quick to administer and do not require clinically trained interviewers. Symptom counts (total scores on the measures) can be used as continuous measures of symptom presence and severity. These measures do not lead to a formal diagnosis, although categorisation above/below a cut-off score can be used to define caseness. Although there is evidence that measures designed in high-income western settings can be used in other cultures, there needs to be a careful process of translation, adaptation and evaluation of reliability, validity and equivalence to the original version [5]. This process requires attention to multiple aspects of the measure: face validity (the subjective impression that the tool as a whole is measuring what it claims to, and that this is a recognized concept in the general population within the cultural context); content validity (whether the included items cover all aspects of the condition, but not aspects that do not form part of the condition; and whether the items are relevant and understood in the context); semantic equivalence (the extent that the original meaning of each item is maintained in the adapted version); technical equivalence (the extent to which the original mode of administration is maintained or whether there are adaptations to make it more usable and understandable in the new context); construct validity (whether the tool measures a coherent state/condition evaluated quantitatively through: (1) measurement of internal consistency, (2) correlation with measures of related constructs (convergent validity), and (3) factor structure); and criterion validity (how the tool performs against an accepted reference “gold standard” measure) [6, 7].

For the Generation Malawi and Healthy Lives Malawi studies, the 7-item Generalised Anxiety Disorder scale (GAD-7), the 9-item Patient Health Questionnaire (PHQ-9); and the ultra-brief Primary Care PTSD Screen for DSM-5 (PC-PTSD-5) were chosen as measures of anxiety, depression and post-traumatic stress disorder respectively. The PHQ-9 and GAD-7 are widely used [8, 9] and have been included by Wellcome in a suite of “common metrics” to enhance comparability of results across studies [10]. The brevity of the PC-PTSD-5 makes it suitable for use in a long questionnaire covering multiple domains administered in community and clinic settings.

The GAD-7 has been validated in a number of countries in Sub-Saharan Africa (SSA): Kenya [11, 12], South Africa [13–17], Ethiopia [18], Ghana [19], Côte d’Ivoire [19], Zimbabwe [20], Mozambique [21]; 4 of these studies have included criterion validation [13, 16, 20, 21]. In Malawi, it has been translated into the Chichewa and Chitumbuka languages and used in a study of perinatal women [22] but has not been formally validated. The PHQ-9 has been validated in 39 studies in SSA, either in English or translation [12–16, 20, 23–55], 21 of which include criterion validation. In Malawi, a Chichewa version was adapted and validated for use in non-communicable disorders (NCD) clinic attenders [40]. Although it has been translated into Chitumbuka [56], this version has not been formally validated. The PC-PTSD-5 has been validated in Kenya (Swahili version) [57], Mozambique (Portuguese version) [58] and in 2 studies in South Africa (English and Ixhosa versions) [15, 16]. It has not been validated in Malawi.

In this study, conducted as formative work for the Generation Malawi and Healthy Lives Malawi studies, we set out to translate and adapt the GAD-7, PHQ-9 and PC-PTSD-5 into the most widely spoken languages in Malawi (Chichewa and Chitumbuka) and evaluate the psychometric properties of the measures with regard to face, content, construct and criterion validity.

## Methods

### Study context

Malawi is a low-income country in south-eastern Africa with a population of 21.7 million [59]. It is 172nd out of 193 countries on the 2022 UNDP Human Development Index [60]. The two study locations are Health and Demographic Surveillance System (HDSS) sites: an urban, predominantly Chichewa speaking, HDSS site (population ≈ 53000) in Area 25 Lilongwe (the capital city of Malawi), and a rural, predominantly Chitumbuka speaking, HDSS site (population ≈ 65000) in the southern part of Karonga District in northern Malawi [3].

The Healthy Lives Malawi study (led by the Malawi Epidemiology and Research Unit (MEIRU)) is an intergenerational longitudinal population study of chronic conditions focussed on understanding how socioeconomic, psychological, biological, nutritional, and infectious factors affect families’ health and wellbeing over time. Healthy Lives Malawi is collecting cross sectional data on 30,000 adults [3]. The linked Generation Malawi family and birth cohort is recruiting 5000 pregnant women (and their spouses) and following them and their offspring up to 3 years postnatally.

### Ethical approval

The study was approved by the University of Malawi, College of Medicine Research and Ethics Committee (COMREC) (P.11/19/2865) and the University of Glasgow, College of Medical, Veterinary & Life Sciences Ethics Committee (200190183).

### Mental Health Measures

#### a. Anxiety

The 7-item Generalized Anxiety Disorder Scale (GAD-7) enquires about the frequency in the last 2 weeks of 7 core symptoms of Diagnostic and Statistical Manual (DSM) generalised anxiety disorder [61]. Each item is scored from 0-3 and the maximum total score is 21.

#### b. Depression

The 9-item Patient Health Questionnaire (PHQ-9) enquires about the frequency in the last 2 weeks of 9 core symptoms of DSM major depressive episode [4]. Each item is scored from 0-3 and the maximum total score is 27. It also includes an additional item on functional impairment scored separately (0-3) from the main scale.

#### c. Post-Traumatic Stress Disorder

The Primary Care PTSD Screen for DSM-5 (PC-PTSD-5) is a 5-item ultra-brief screening tool designed to identify individuals with probable PTSD in primary care settings [62]. A gating question lists various “unusually or especially frightening, horrible, or traumatic” events and asks whether the respondent has ever experienced one of these. Respondents who report experiencing a traumatic event then answer 5 yes/no questions about PTSD symptoms in the past 1 month. Total scores range from 0 (no traumatic event <u>or</u> traumatic event but no symptoms) to a maximum of 5.

#### d. Somatic complaints

People in distress may present to health services with non-specific somatic complaints such as headache, general body pains and gastrointestinal symptoms. The inclusion of somatic items as part of screening for common mental health conditions may improve detection rates [63]. We therefore added 3 items to the battery of mental health screening measures, chosen from the PHQ-15 [64]. These items are scored similarly to GAD-7 and PHQ-9 (0-3) and we calculated a sum score (0-9).

### Study phases

The study was conducted in 2 phases. In phase 1, we undertook translation, adaptation, expert review, cognitive interviewing, and piloting to produce final versions of the measures in both languages (Chichewa and Chitumbuka). We assessed (and attempted to maximise) face and content validity, and the semantic and technical equivalence to the original English versions. In phase 2 we conducted criterion validation of the GAD-7, PHQ-9 and PC-PTSD-5 against Diagnostic and Statistical Manual version 5 (DSM-5) diagnoses of generalised anxiety disorder, major/minor depressive episode, and PTSD respectively. We assessed construct validity (internal consistency, convergent validity and factor structure) and evaluated measurement invariance between the 2 language versions of each measure. Finally, we estimated weighted prevalences of the DSM-5 diagnoses.

### Phase 1

A trilingual researcher (medical graduate) undertook initial forward translation of the measures. Through a literature search we identified any existing Chichewa and Chitumbuka translations of the tools [22, 40, 56] and compared these to our initial translations. We held translation consensus meetings of research staff, fieldworkers and an expert panel. The translated PHQ-9 and GAD-7 were back-translated by a trilingual non-medical professional into English. These back translations were scrutinised and further changes made. We conducted 2 FGDs in each site with 4 community participants per group in which participants were asked about their understanding of the items, and whether they recognised the concepts in their context. The participants were then administered the measures individually (in a private setting) and invited to describe their understanding of each item, and their choice of response (“cognitive interviewing” [65]).

Regarding the PTSD measure, we had originally planned to use the longer PTSD Checklist for DSM-5 (PCL-5) [66] and it was this measure that was discussed in expert and fieldworker consensus meetings, and in 2 FGDs with community members. However, it became evident that the PCL-5 was too long to include in the questionnaire battery for the GM and HLM studies. We decided to change to use the much shorter PC-PTSD-5. The translations of the relevant items in PCL5 were adapted to the PC-PTSD-5 format; these were discussed with fieldworkers and piloted. However, back translation, further FGDs and cognitive interviewing were not conducted for the measure.

We developed a visual response tool for GAD-7 and PHQ-9 to improve comprehension as previous research had found that respondents find it difficult to remember 4 response options unless these are read out after each item, or a visual representation of the options is used [67, 68]. We scrutinised various visual aid designs to represent the levels of severity/frequency from Malawi [56, 68, 69], UK [70] and Nepal [71]. We discussed the options within the research team and with a group of fieldworkers, and then piloted the aid. To achieve consistency in the explanations that fieldworkers should give if participants asked for clarification, we developed a laminated one-page sheet of standardised explanations. Such an approach had been used previously in validation of the Self Reporting Questionnaire (SRQ-20) in Malawi [72].

The measures were loaded onto tablets using Open Data Kit (ODK). In each site, two experienced MEIRU field interviewers were trained to administer the questionnaires. The training included observed pilot administration of the measures with feedback given to the fieldworkers to ensure fidelity to protocol. We did not conduct formal inter-rater reliability.

Any remaining issues regarding translation were resolved during this piloting, and final versions of the measures in each language were agreed. The field interviewers were also trained in consent procedures and collection of basic demographic information (age, sex, highest educational level).

### Phase 2

We recruited participants between 14 January and 21 May 2021 from facilities and community settings in the Lilongwe and Chilumba DHSS sites. We included participants who were 18 years and above on the day of recruitment, and able to give informed consent. Each participant was invited to read the information sheet and consent form. For participants who were unable to read or write, the Information sheet and consent form were read out in the presence of an impartial witness. Written informed consent was obtained where possible. Where a participant was willing to provide consent but unable to write, verbal consent was documented. Participants were then asked to sign the consent form using a signature or thumb print and to indicate their name and date. Where an impartial witness was involved, they were requested to sign and date the consent form. The field interviewer also co-signed the form. One copy of the signed consent form was provided to the participant, while another copy was retained by the interviewer for study records. In each site, we recruited convenience samples of the following 3 groups:

- pregnant mothers attending antenatal clinics at Area 25 Health Centre in Lilongwe and Chilumba Rural Hospital in Karonga;
- postnatal mothers attending for infant vaccination or growth monitoring services at the under-five clinics in the same settings;
- men from the community surrounding the two facilities.

### Reference standard diagnostic interviews

The reference standard criteria were diagnoses of Generalised Anxiety Disorder (GAD), current major or minor Depressive Episode, and PTSD made using the Structured Clinical Interview for DSM version 5 (SCID-5) [73]. The interviewer team comprised 5 mental health clinical officers (clinicians with a Diploma in clinical medicine and 2-year BSc in clinical mental health). They translated the SCID modules into Chitumbuka and Chichewa, and agreed final versions after discussion. Training was led by JM, an experienced SCID interviewer. Initial interviews were conducted with one interviewer administering the SCID and the other(s) observing (on a rotating basis). In Karonga, the 2 interviewers saw 12 patients together and had 100% agreement in the diagnoses made. In Lilongwe, the 3 interviewers saw 4 patients together with 100% agreement for GAD and PTSD, and 92% agreement for depression. Throughout the study, the interviewers in Karonga (JM, MMa) and Lilongwe (JM, MMs, TM) discussed every case with their colleague(s) and agreed consensus diagnoses.

### Sampling of screened participants for SCID interview

To maximise efficiency of the study, we invited a weighted sample of screened participants to have SCID interview and adjusted for this weighting in the statistical analyses [74]. For the weighted sampling, we defined a “high scorer” as a participant who fulfilled <u>any</u> of the following criteria: GAD-7 total score ≥ 3, PHQ-9 total score ≥ 5, a score of ≥ 1 on the PHQ-9 suicide item, or reporting a traumatic life event on the PC-PTSD-5. All “high scorers” were invited to have SCID interview. A random sample of approximately every third “low scorer” was also invited. Of those invited for SCID (either “high scorers” or “low scorers”), some refused and there were some who were missed or lost for other reasons.

### Safety considerations

Participants who were assessed during the SCID interview to have significant distress were provided with brief unstructured counselling by the interviewer (mental health clinical officer). In addition, those assessed as requiring ongoing intervention for management of a mental health condition and/or suicide risk were referred to local mental health services. Participants identified as being at risk of intimate partner violence were signposted to appropriate support.

### Statistical analysis

All analyses were conducted separately for the Chitumbuka and Chichewa versions of the measure. We calculated descriptive statistics for sample demographic characteristics, statistical analyses of construct and criterion validity, and estimates of prevalence of GAD, Major and Minor Depressive Episode and PTSD. Analyses were performed using R version 4.3.1 using the following packages: tidyverse, dplyr, ltm, ggplot2, psych, nFactors, corrplot, Hmisc, and lavaan.

#### a. Construct validity

Construct validity of the GAD-7, PHQ-9 and PC-PTSD-5 was assessed as follows: for internal consistency, we calculated Cronbach’s alpha for the whole scale in each measure (>0.7 interpreted as acceptable/good); for convergent validity, we calculated Spearman’s correlations (for non-parametric data) between GAD-7 and PHQ-9, GAD-7 and PC-PTSD-5, and PHQ-9 and PC-PTSD-5 total scores (for the 2 latter analyses, participants with no trauma on PC-PTSD-5 were excluded); and for factor structure, we conducted confirmatory factor analysis (CFA). CFA was conducted using lavaan package in R. The number of factors in the CFA models was based on previous studies (identified through literature review) and we tested alternative models (e.g., a one-factor model vs. a two-factor model) to find the structure that best represented the data. For CFA of PC-PTSD-5, participants with no trauma were excluded.

Before conducting CFA, for each measure, we assessed inter-item correlations using polychoric correlation and assessed sample size adequacy of the data for factor analysis using Kaiser-Meyer-Olkin (KMO) test and sample suitability using Bartlett’s test for sphericity. The Comparative Fit Index (CFI) and the Tucker–Lewis Index (TLI) were used to determine whether the model fit the data better than a more restricted baseline model. The root mean square error of approximation (RMSEA) was used to measure how closely the model fit the data. In addition, using multi-group CFA, we conducted measurement invariance tests to assess if the mental health measures had the same measurement properties in Chichewa and Chitumbuka study populations. Tests for measurement invariance consist of a series of model comparisons whereby several parameters can be constrained across groups [75]. First is a configural invariance model whereby the same factor structure is imposed on all groups to test whether the basic factor structure is consistent between groups; second is metric/weak invariance model in which the factor loadings are constrained to be identical across groups to test whether there is non-uniform item bias; and the third is a scalar/strong invariance model in which the item factor loadings and intercepts are constrained to be identical across groups to test whether there is uniform item bias present in an item. The model comparison test first compares the configural model versus the metric model, while the second test compares the metric model versus the scalar model. At each step of the series, the constrained model is assessed to see if it improves or worsens the model fit to the data by evaluating fit statistics (Chi-square, CFI, RMSEA and SMR) and change in these fit statistics. In the series of tests for invariance, invariance was judged to hold if changes in TLI and CFI values were ≤0.01; change in RMSEA value was ≤0.015; change in SRMR value was ≤0.033 in metric and ≤0.01 in the scalar model, and change in chi-square p-value was >0.05.

#### b. Criterion validity

We calculated the test characteristics of GAD-7, PHQ-9, PC-PTSD-5 against reference diagnoses of GAD, major/minor depressive episode and PTSD respectively. Test characteristics (sensitivity, specificity, positive predictive value (PPV), negative predictive value (NPV)) and Youden’s index for each cut-off score were calculated. For the PHQ-9, we also investigated whether adding the 3-item somatic symptoms total score to the PHQ-9 total score improved identification of major/minor depressive episodes.

Inverse probability sampling weights, stratified by study site, were used to account for the different proportions invited for SCID assessment between “high scorers” vs “low scorers”. When calculating the weights, we used the inverse of the number who had the SCID administered divided by the number who received the screening tool. When calculating weights, we assumed no difference between those who had the SCID and those who were invited but who refused or were otherwise missed/lost. We conducted a sensitivity analysis excluding all data on participants who refused SCID or who were missed/lost; this made minimal difference to the area under the Receiver Operating Characteristic (ROC) curve (AUC) or prevalence estimates.

ROC curves for the measures in the 2 languages were obtained by plotting sensitivity against 1-specificity for each possible cut-off score. Weighted AUC estimates were obtained with 95% confidence intervals calculated using nonparametric bootstrap with 1000 re-samples to account for the two-phase study design and inverse probability weighting.

#### c. Prevalence

Weighted estimates of prevalence of SCID-diagnosed GAD, major/minor depressive episode and PTSD for both sites were calculated using inverse sample weights, recognising that these were convenience samples and not necessarily representative of the population from which they were drawn.

## RESULTS

### Phase 1

#### a. Face and content validity

Field workers and community FGD participants recognized local distress states consistent with diagnostic concepts of depression and generalised anxiety, and agreed that the PHQ-9 and GAD-7 included items found in these conditions. We did not investigate whether the measures mapped perfectly onto local concepts. Regarding the PC-PTSD-5, participants recognised the concept of a distress state that can follow a life-threatening event, although they found some of the items difficult to understand and translate, e.g., “Felt numb or detached from people, activities, or your surroundings?”.

#### b. Semantic equivalence

For each measure, final translations were agreed that maintained, as far as possible, the meaning of the original items and reflected language usage in the study areas. In Karonga, the spoken Chitumbuka differs from that spoken in other parts of the northern region (that had been captured in initial translations), so modifications were made to the translations to reflect this.

#### c. Technical Equivalence

To improve acceptability and comprehension, we made several adaptations that affected technical equivalence to the original measures:

1. The original measures were self-report, designed to be completed on paper or electronically by the participant. To be consistent with overall study procedures, and avoid excluding participants with low literacy, we changed items from statement to question format.
2. We found that participants tended to forget the time period referred to in the measures so “In the last 2 weeks” was added to each item in the PHQ-9 and GAD-7.
3. The original PHQ-9 and GAD-7 use 4 response options to ask *how often* the person has the symptom in the last 2 weeks (not at all, several days, more than half the days, nearly every day). Some participants found this difficult to comprehend, creating uncertainty about which response to give. We therefore simplified the prompt to “how much” and the responses to 1-2 words that were simple but that we felt captured the essence of the original (Not at all, Sometimes, Usually, Almost Always). This also made the measure more user-friendly for field interviewers.
4. As described above, we developed a visual aid that included the response item text but also visual representations. We adapted a prompt used in Nepal [71] where the responses were represented by glasses with differing amounts of water in them. These were used in this validation study. However, in more prolonged use in the main study, it was felt that the explanation about the aid was long, and some participants found it confusing as the increasing fullness of the glass represented more severe symptoms, whereas having a full glass of water could be seen as positive. Therefore, it was changed to being optional for fieldworkers to deliver, as long as they read out the 4 responses after each item.
5. Following a procedure previously used in adaptation of the SRQ-20 [72] we provided a script with examples to be read verbatim by the fieldworker if a person did not understand the question. This limited the potential for interviewers to give their own interpretations when explaining the questions.
6. In the PHQ-9, the psychomotor agitation/retardation question (Q8) was difficult to understand so we split it into 2 sub-items (Q8a and Q8b), asking first about retardation and then agitation. To maintain the original total score (max 27), only the <u>highest</u> of these 2 sub-items is included in the total score (or only 8a score if Q8a and Q8b equal)
7. In the PC-PTSD-5, we added “a very difficult childbirth when you thought that you or the baby would die” to the types of traumatic events listed in the introduction and we added recording of what type of traumatic event the respondent had experienced.

Final versions of the measures are shown in Supporting Information (**S1 text: GAD-7, PHQ-9, Somatic Complaints, PC-PTSD-5 modified English, Chitumbuka and Chichewa versions.)**

### Phase 2

#### Characteristics of study participants

Eight hundred participants were administered the screening tools: n=342 in the rural Chitumbuka-speaking site (Karonga), and n=458 in the urban Chichewa-speaking site (Lilongwe). Participant characteristics are shown in **Table 1**.

**Table 1:** Descriptive characteristics of all screened participants.

|  | All | Karonga<br>(Chitumbuka-speaking) | Lilongwe<br>(Chichewa-speaking) |
| --- | --- | --- | --- |
| **Characteristic** | N = 800 | N = 342 | N = 458 |
| **Age (years)** |  |  |  |
| Mean (SD) | 29.5 (11.3) | 30.8 (12.7) | 28.5 (10.0) |
| Median (Q1, Q3) | 26.0 (21.0, 33.0) | 26.5 (21.0, 36.0) | 26.0 (21.0, 32.0) |
| **Sex** |  |  |  |
| Female postnatal | 134 (16.8%) | 75 (21.9%) | 59 (12.9%) |
| Female pregnant | 271 (33.9%) | 91 (26.6%) | 180 (39.3%) |
| Male | 395 (49.4%) | 176 (51.5%) | 219 (47.8%) |
| **Interview language** |  |  |  |
| Chitumbuka | 342 (42.8%) | 342 (100.0%) | 0 (0.0%) |
| Chichewa | 458 (57.3%) | 0 (0.0%) | 458 (100.0%) |
| **Interview location** |  |  |  |
| In facility (e.g. ANC, vaccination clinic) | 423 (52.9%) | 177 (51.8%) | 246 (53.7%) |
| At home | 371 (46.4%) | 165 (48.2%) | 206 (45.0%) |
| Other | 6 (0.8%) | 0 (0.0%) | 6 (1.3%) |
| **Marital status** |  |  |  |
| Never married | 176 (22.0%) | 67 (19.6%) | 109 (23.8%) |
| Married | 557 (69.6%) | 237 (69.3%) | 320 (69.9%) |
| Divorced/separated | 60 (7.5%) | 35 (10.2%) | 25 (5.5%) |
| Widowed | 7 (0.9%) | 3 (0.9%) | 4 (0.9%) |
| **Education** |  |  |  |
| None | 11 (1.4%) | 1 (0.3%) | 10 (2.2%) |
| Primary | 320 (40.0%) | 175 (51.2%) | 145 (31.7%) |
| Secondary | 422 (52.8%) | 156 (45.6%) | 266 (58.1%) |
| Higher | 47 (5.9%) | 10 (2.9%) | 37 (8.1%) |
| **Employment status** |  |  |  |
| Subsistence farmer | 191 (23.9%) | 182 (53.2%) | 9 (2.0%) |
| “Housewife” | 144 (18.0%) | 16 (4.7%) | 128 (27.9%) |
| Salaried employment | 68 (8.5%) | 19 (5.6%) | 49 (10.7%) |
| Piece work | 152 (19.0%) | 34 (9.9%) | 118 (25.8%) |
| Own business | 142 (17.8%) | 69 (20.2%) | 73 (15.9%) |
| Student | 56 (7.0%) | 11 (3.2%) | 45 (9.8%) |
| Other | 47 (5.9%) | 11 (3.2%) | 36 (7.9%) |

ANC – Antenatal Clinic

#### Screening measure scores

Screening measure scores are shown in in **Table 2**. For the GAD-7, PHQ-9, PHQ-9 functioning item and 3-item somatic complaint measure, total scores (mean (SD) and median (IQR)) are shown. For PC-PTSD-5, proportions of respondents who had experienced any lifetime trauma (and different trauma types) are presented alongside PC-PTSD-5 total score (mean (SD) and median (IQR).

**Table 2.**
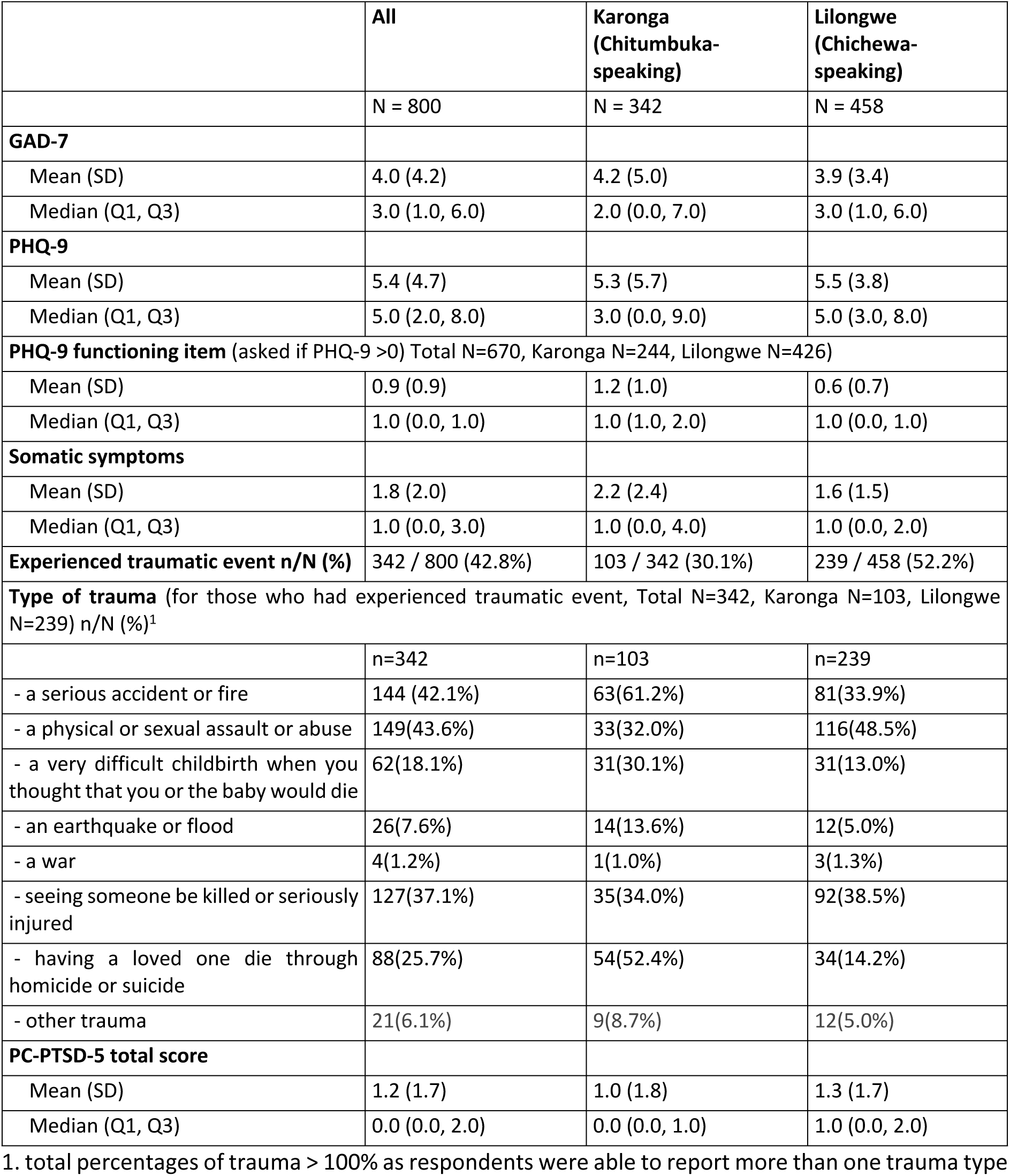
Mental health screening measure scores.

|  | All | Karonga<br>(Chitumbuka-speaking) | Lilongwe<br>(Chichewa-speaking) |
| --- | --- | --- | --- |
|  | N = 800 | N = 342 | N = 458 |
| **GAD-7** |  |  |  |
| Mean (SD) | 4.0 (4.2) | 4.2 (5.0) | 3.9 (3.4) |
| Median (Q1, Q3) | 3.0 (1.0, 6.0) | 2.0 (0.0, 7.0) | 3.0 (1.0, 6.0) |
| **PHQ-9** |  |  |  |
| Mean (SD) | 5.4 (4.7) | 5.3 (5.7) | 5.5 (3.8) |
| Median (Q1, Q3) | 5.0 (2.0, 8.0) | 3.0 (0.0, 9.0) | 5.0 (3.0, 8.0) |
| **PHQ-9 functioning item** (asked if PHQ-9 >0) Total N=670, Karonga N=244, Lilongwe N=426) |  |  |  |
| Mean (SD) | 0.9 (0.9) | 1.2 (1.0) | 0.6 (0.7) |
| Median (Q1, Q3) | 1.0 (0.0, 1.0) | 1.0 (1.0, 2.0) | 1.0 (0.0, 1.0) |
| **Somatic symptoms** |  |  |  |
| Mean (SD) | 1.8 (2.0) | 2.2 (2.4) | 1.6 (1.5) |
| Median (Q1, Q3) | 1.0 (0.0, 3.0) | 1.0 (0.0, 4.0) | 1.0 (0.0, 2.0) |
| **Experienced traumatic event n/N (%)** | 342 / 800 (42.8%) | 103 / 342 (30.1%) | 239 / 458 (52.2%) |
| **Type of trauma** (for those who had experienced traumatic event, Total N=342, Karonga N=103, Lilongwe N=239) n/N (%)<sup>1</sup> |  |  |  |
|  | n=342 | n=103 | n=239 |
| - a serious accident or fire | 144 (42.1%) | 63(61.2%) | 81(33.9%) |
| - a physical or sexual assault or abuse | 149(43.6%) | 33(32.0%) | 116(48.5%) |
| - a very difficult childbirth when you thought that you or the baby would die | 62(18.1%) | 31(30.1%) | 31(13.0%) |
| - an earthquake or flood | 26(7.6%) | 14(13.6%) | 12(5.0%) |
| - a war | 4(1.2%) | 1(1.0%) | 3(1.3%) |
| - seeing someone be killed or seriously injured | 127(37.1%) | 35(34.0%) | 92(38.5%) |
| - having a loved one die through homicide or suicide | 88(25.7%) | 54(52.4%) | 34(14.2%) |
| - other trauma | 21(6.1%) | 9(8.7%) | 12(5.0%) |
| **PC-PTSD-5 total score** |  |  |  |
| Mean (SD) | 1.2 (1.7) | 1.0 (1.8) | 1.3 (1.7) |
| Median (Q1, Q3) | 0.0 (0.0, 2.0) | 0.0 (0.0, 1.0) | 1.0 (0.0, 2.0) |

1. total percentages of trauma > 100% as respondents were able to report more than one trauma type

#### Participant flow to reference diagnostic interviews

Of the n=800 screened, n=470 had a SCID interview. Using the sampling procedures for “high scorers” and “low scorers”, in Karonga, 257/342 (75%) participants were invited for SCID interview of whom n=219/257 (85%) were interviewed and the rest refused or were missed/lost. In Lilongwe n=411/458 (90%) were invited for the SCID interview of whom 251/411 (61%) had the SCID interview and the rest refused or were missed/lost. **Table 3** shows the number of “high scorers” and “low scorers” who were invited for SCID, those who refused or were missed/lost for other reasons, and those who had a SCID interview completed. The calculated inverse probability weights are shown, along with the absolute numbers of those participants diagnosed with the relevant reference condition.

**Table 3.**
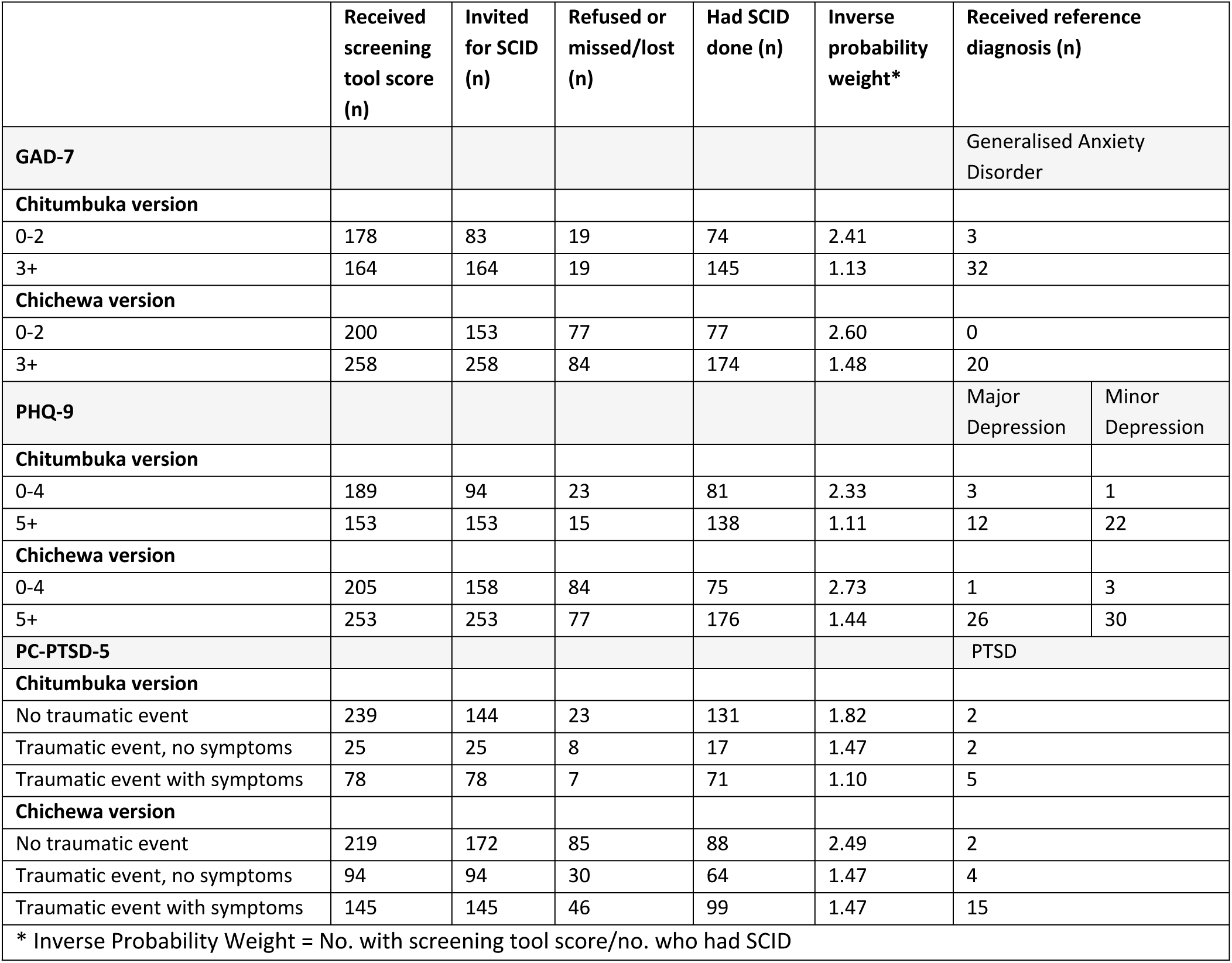
Inverse probability sampling weights by measure and study site/language.

#### Prevalence of DSM-5 diagnoses

Weighted prevalence estimates of DSM-5 Generalized Anxiety Disorder, Major and Minor depressive episode and PTSD in the whole sample and the 2 study sites are shown in **Table 4**.

**Table 4.** Weighted Prevalence Estimates of DSM-5 Generalized Anxiety Disorder, Major and Minor Depressive Episode, and PTSD.

|  | All<br>% (95% CI) | Karonga (Chitumbuka-speaking)<br>% (95% CI) | Lilongwe (Chichewa-speaking)<br>% (95% CI) |
| --- | --- | --- | --- |
| **Generalized Anxiety Disorder** | 9.5% (7.7% - 11.8%) | 12.7% (9.6% - 16.6%) | 6.5% (4.6% - 9.1%). |
| **Major depression** | 6.4% (4.9% - 8.4%) | 5.9% (3.9% - 9.0%) | 8.8% (6.5% - 11.7%). |
| **Minor depression** | 8.3% (6.7% - 10.5%) | 7.8% (5.4% - 11.2%) | 11.3% (8.7% - 14.6%) |
| **Minor or Major Depression** | 14.8% (12.5% - 17.5%) | 13.8% (10.5% - 17.8%). | 19.9% (16.6% - 23.9%). |
| **PTSD** | 5.4% (4.1% - 7.2%) | 3.5% (2.0% - 6.1%) | 7.2% (5.1% - 9.9%) |

#### Consstruct and Criterion Validity

##### GAD-7

###### a. Internal consistency

Cronbach’s alpha: Chitumbuka version = 0.892 (95% CI: 0.862 - 0.914); Chichewa version = 0.804 (95% CI: 0.773 - 0.832).

###### b. Convergent validity

GAD-7 total score was highly correlated with PHQ-9 total score in both versions (Chitumbuka r=0.80; Chichewa r=0.78; both p-value <0.05). GAD-7 score correlation with PC-PTSD-5 score (for participants who experienced trauma) was weakly positive (r=0.39; p-value <0.05) in Chitumbuka, and moderately positive in Chichewa (r=0.46; p-value <0.05).

###### c. Factor analysis

Correlation matrices are shown in Supporting Information **S2A Table** and **S2B Table**. The KMO test statistics indicated that the sampling for the GAD-7 items and construct was adequate for factor analysis for both versions. The GAD-7 construct had an overall measure of sampling adequacy (MSA) value of 0.91 in Chitumbuka and 0.89 in Chichewa, with all individual item MSA values ≥0.86 for both versions. Bartlett’s test for sphericity (Chitumbuka, Chi-square = 1162.18, Df = 21, p-value <0.005; Chichewa, Chi-square = 764.29, Df = 21, p-value <0.005) implied that there was no problem of singularity in the measured GAD-7 item scores hence making both versions suitable for factor analysis.

The CFA analysis based on a one factor model indicated that all GAD-7 items loaded onto the one-factor structure in both Chitumbuka and Chichewa. Fit indices were good for both the Chitumbuka version (RMSEA = 0.07 [90% CI 0.05 - 0.01], SRMR = 0.029; CFI = 0.97; TLI = 0.96) and Chichewa version (RMSEA = 0.048 [90% CI 0.02 - 0.07], SRMR= 0.03; CFI = 0.98; TLI = 0.97).

###### d. Measurement invariance

The tests of measurement invariance between the 2 language versions of the GAD-7 are reported in Supporting Information **S4 Table**. For the baseline configural model for GAD-7, the underlying factor structure was comparable across the groups and fit statistics indicated a good fit (RMSEA=0.062; CFI=0.978; TLI=0.967; SRMR=0.02). For the metric model, compared with the configural model, the change in chi-square p-value (Pr(ΔΧ^2^)<0.001) and change in CFI (ΔCFI=0.016) indicated that the metric model worsened the fit, and the changes in RMSEA (ΔRMSEA=0.01), TLI (ΔTLI=0.01) and SRMR (ΔSRMR=0.33) were on the borderline of cutoffs, indicative of invariance violation. The scalar model performed worse than the baseline model (Pr(ΔΧ^2^)<0.001; ΔCFI=0.034; ΔRMSEA=0.02; ΔSRMR=0.011) indicating that scalar invariance did not hold.

###### 2. Criterion validity

Sensitivity, specificity, positive and negative predictive values at each cut-off score on the Chitumbuka and Chichewa GAD-7 versions are shown in Supporting Information **S5A table**. ROC curves are shown in **Figure 1**. The AUC for detection of generalised anxiety disorder in Chitumbuka was 0.759 (95%CI: 0.634, 0.871) and in Chichewa was 0.868 (95%CI: 0.812, 0.915). The cut-off with the highest Youden index was 5/6 in both Chitumbuka (sensitivity 70.4%. specificity 74.6%, Youden Index 0.45) and Chichewa (sensitivity 90%, specificity 73%, Youden Index 0.63) (Supporting Information **S5A table**).

**Figure 1:**
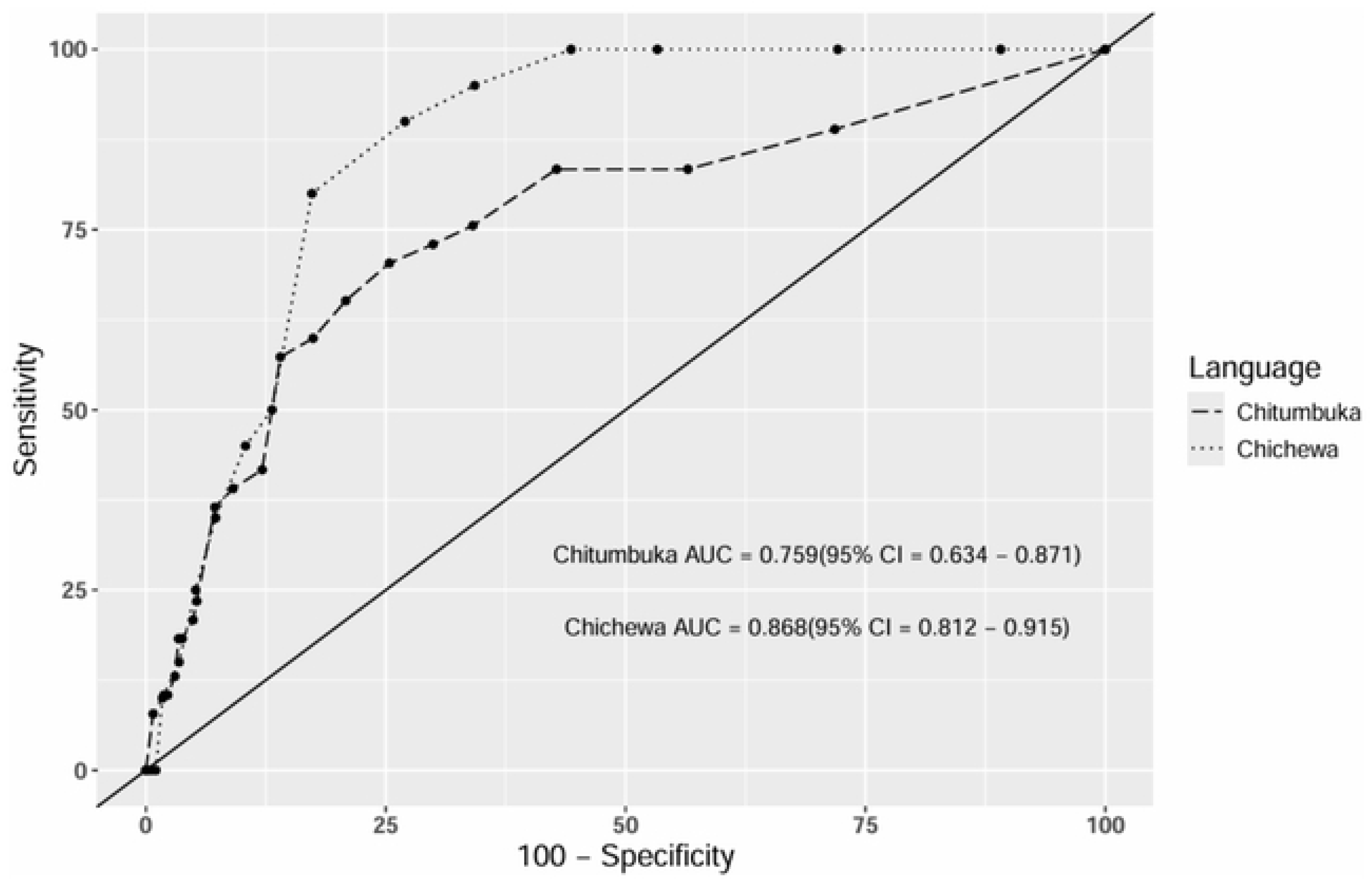
ROC curves for GAD-7 vs Generalised Anxiety Disorder reference diagnosis in Chitumbuka and Chichewa.

##### PHQ-9

###### a. Internal consistency

Cronbach’s alpha: Chitumbuka version = 0.865 (95%CI: 0.837, 0.887); Chichewa version = 0.749 (95% CI: 0.717 - 0.781).

###### b. Convergent validity

Correlations of PHQ-9 and GAD-7 scores are reported above. PHQ-9 score correlation with PC-PTSD-5 score (for participants who experienced trauma) was weakly positive in Chitumbuka (r=0.38; p-value <0.05), and moderately positive in Chichewa (r=0.43; p-value <0.05).

###### c. Factor analysis

Correlation matrices are shown in Supporting Information **S2C Table** and **S2D Table**. The KMO test statistics indicated that the sampling for the PHQ-9 items and construct was adequate, thus appropriate for factor analysis in both versions. The PHQ-9 construct had an overall measure of sampling adequacy (MSA) value of 0.9 in Chitumbuka and 0.86 in Chichewa, with all individuals item MSA values ≥0.85 in Chitumbuka and ≥0.82 in Chichewa. Bartlett’s test (Chitumbuka, Chi-square = 1103.46, Df = 36, p-value <0.005; Chichewa, Chi-square = 648.68, Df = 36, p-value <0.005) indicated no singularity issues.

CFA based on a one factor Chitumbuka model indicated that the data fit the model marginally on this one-factor structure (RMSEA = 0.101 [90% CI 0.084, 0.120], SRMR = 0.051; CFI = 0.912; TLI = 0.883). A two factor CFA model indicated a reasonable fit to the data (RMSEA = 0.083 [90% CI 0.065, 0.103], SRMR = 0.043; CFI = 0.943; TLI = 0.921). In Chichewa, a two factor CFA model in Chichewa had a good fit to the data (RMSEA = 0.035 [90% CI 0.01, 0.055], SRMR= 0.035; CFI = 0.976; TLI = 0.967).

###### d. Measurement invariance

Tests of measurement invariance between the 2 language versions of the PHQ-9 are reported in Supporting Information **S3 Table**. For the baseline configural model for PHQ-9, the underlying factor structure was comparable across the groups with an acceptable fit (RMSEA=0.084; CFI=0.911; SRMR=0.05). The change in chi-square probability indicated the metric model fit significantly worsened compared to the baseline configural model (Pr(ΔΧ^2^)=0.002), however the change in CFI (ΔCFI=0.01) was on the borderline of recommended cutoffs, while the changes in RMSEA, TLI and SRMR were below the recommended cutoffs (ΔRMSEA <0.002; ΔTLI=0.004; ΔSRMR=0.014) suggesting possible support for metric invariance. The scalar model significantly worsened the fit compared to the metric model (Pr(ΔΧ^2^) <0.001; ΔCFI=0.065, ΔTLI=0.053, ΔRMSEA =0.017, ΔSRMR=0.014) indicating that scalar invariance did not hold.

###### 2. Criterion validity

Sensitivity, specificity, positive and negative predictive values for each of the possible cut-off scores on the PHQ-9 Chitumbuka and Chichewa versions for identifying DSM-5 major depression and minor-or-major depression are shown in Supporting Information **S4B table** and **S4C table**.

###### a. Major depression

ROC curves for major depression are shown in **Figure 2a**. The AUC for identifying major depression in Chitumbuka was 0.634 (95%CI: 0.441, 0.869) and in Chichewa was 0.843 (95%CI: 0.721, 0.927). The cut-off with the highest Youden index was 8/9 in both Chitumbuka (sensitivity 60%, specificity of 78%, Youden Index 0.38) and Chichewa (sensitivity 79%, specificity of 86%, Youden Index 0.64) (Supporting Information **S4B table**).

**Figure 2a:**
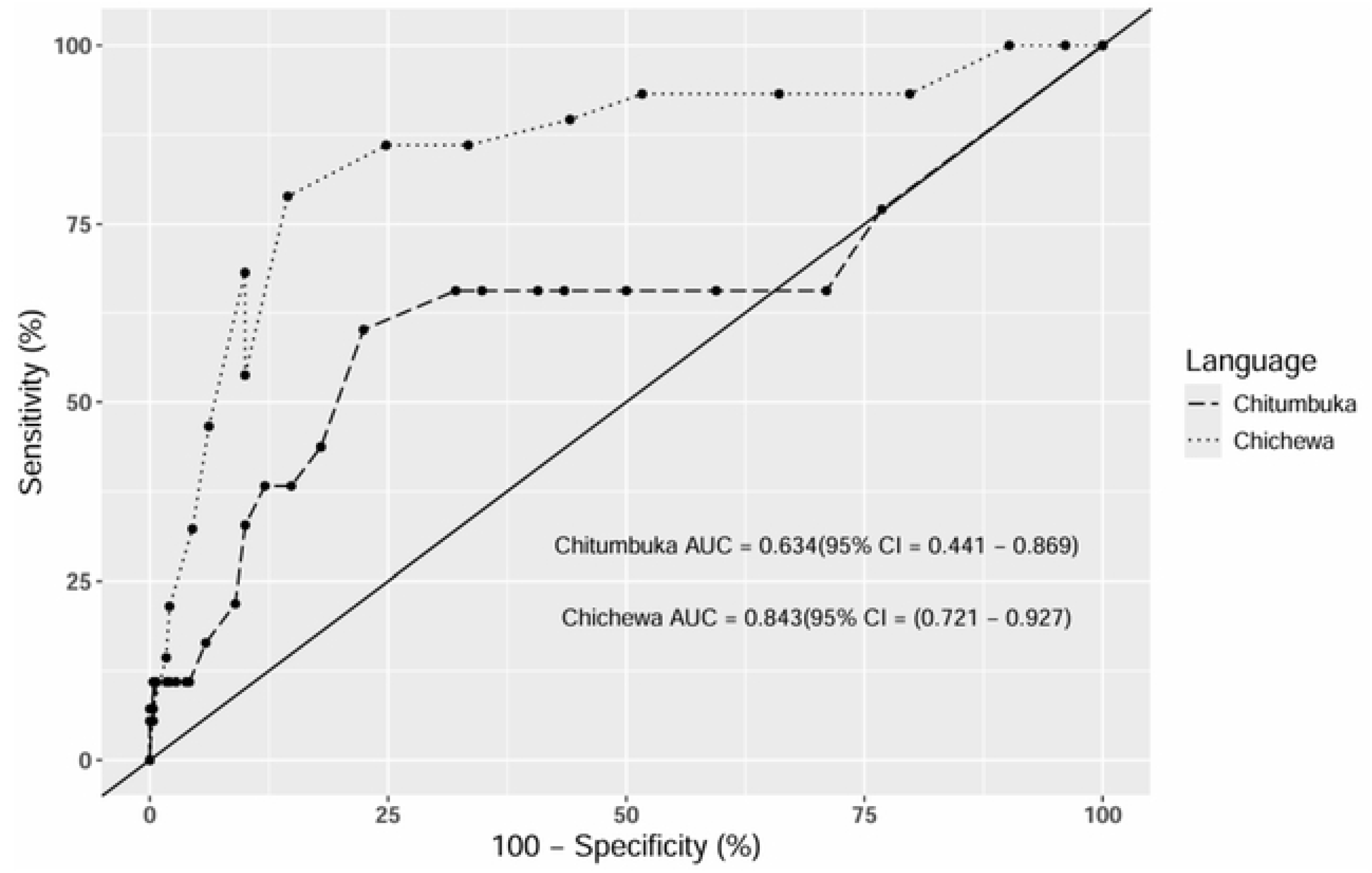
ROC curves for PHQ-9 vs major depressive episode reference diagnosis in Chitumbuka and Chichewa.

**Figure 2b:**
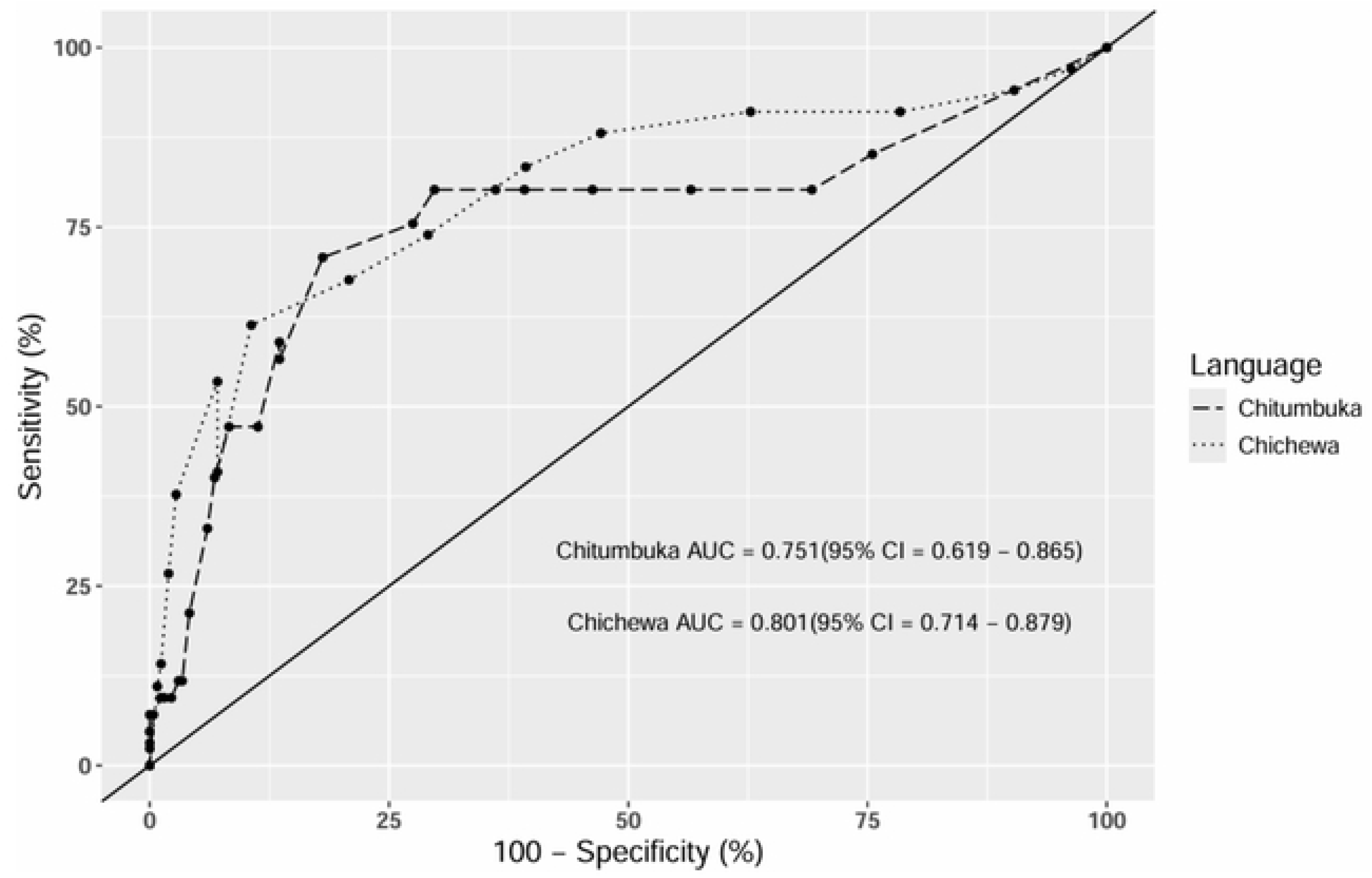
ROC curves for PHQ-9 vs major-or-minor depressive episode reference diagnosis in Chitumbuka and Chichewa.

###### b. Minor-or-major depression

ROC curves for minor-or-major depression are shown in **Figure 2b**. The AUC for identifying minor-or-major depression in Chitumbuka was 0.751 (95%CI: 0.619, 0.865) and in Chichewa was 0.801 (95%CI: 0.714, 0.879). The cut-off with the highest Youden index was 8/9 in both Chitumbuka (sensitivity 71%, specificity 82%, Youden Index 0.53) and Chichewa (sensitivity 61%, specificity 89%, Youden Index 0.51) (Supporting Information **S5C Table**).

###### c. Addition of 3 somatic complaint items

Addition of the 3 additional somatic complaints items to the PHQ-9 (increasing the maximum total score to 36) had a negligible effect on the performance of the measures. The AUC for detecting major depression changed from 0.634 to 0.610 in Chitumbuka and from 0.843 to 0.881 in Chichewa. For detecting minor-or-major depression, the weighted AUC changed from 0.751 to 0.740 in Chitumbuka and from 0.801 to 0.804 in Chichewa.

##### PC-PTSD-5

###### a. Internal consistency

Cronbach’s alpha: Chitumbuka version = 0.769 (95% CI: 0.690, 0.827); Chichewa version = 0.696 (95% CI: 0.625, 0.754).

###### b. Convergent validity

Correlation of PC-PTSD-5 scores with GAD-7 and PHQ-9 scores are reported above.

###### c. Factor analysis

Correlation matrices are shown in Supporting Information **S2E Table** and **S2F Table**. The KMO test statistics indicated that the items in the PC-PTSD-5 construct were correlated, and the dataset was appropriate for a factor analysis for both versions. The PC-PTSD-5 construct had an overall measure of sampling adequacy (MSA) value of 0.74 in both Chitumbuka and Chichewa. Individual items had an MSA value ranging from 0.70-0.86 in Chitumbuka, and 0.68-0.81 in Chichewa. Bartlett’s test for sphericity (Chitumbuka, Chi-square = 124.58, Df = 10 p-value <0.005; Chichewa, Chi-square = 189.14, Df = 10, p-value <0.005) implied that there were no singularity issues in the measured PC-PTSD-5 items scores.

A one factor CFA model indicated that all the PC-PTSD-5 items loaded onto this structure in both Chitumbuka and Chichewa. Fit indices for the unidimensional factorial structure of the PC-PTSD-5 were good in Chitumbuka (RMSEA = 0.06 [90% CI 0.0, 0.16], SRMR= 0.036; CFI = 0.99; TLI = 0.97) and reasonable in Chichewa (RMSEA = 0.087 [90% CI 0.035, 0.14], SRMR= 0.04; CFI = 0.95; TLI = 0.90).

###### d. Measurement invariance

Tests of measurement invariance between the 2 language versions of the PC-PTSD-5 are reported in Supporting Information **S3 Table**. For the baseline configural model for PC-PTSD-5, the underlying factor structure was comparable across the groups; the CFI and TLI indicated a good fit for the configural model while the RMSEA indicated acceptable fit (CFI=0.964; TLI=0.928; RMSEA=0.08; SRMR=0.03). The change in chi-square probability and the change in CFI showed that the metric model significantly worsened the fit (Pr(ΔΧ^2^)=0.04; ΔCFI =0.02) however the changes in RMSEA, TLI and SRMR indicated that the metric model did not worsen the fit (ΔRMSEA <0.004; ΔTLI=0.008; ΔSRMR =0.024); overall, these findings indicate possible invariance violation. The scalar model worsened the fit compared to the metric model (Pr(ΔΧ^2^)<0.001; ΔCFI=0.08, ΔTLI=0.07, ΔRMSEA=0.03, ΔSRMR=0.02) indicating that invariance was not supported.

###### 2. Criterion validity

Sensitivity, specificity, positive and negative predictive values for each of the possible cut-off scores on the PC-PTSD-5 Chitumbuka and Chichewa versions for identifying DSM-5 PTSD are shown in Supporting Information **S4D table**. ROC curves are shown in **Figure 3**. The AUC for detection of PTSD in Chitumbuka was 0.682 (95%CI: 0.519, 0.853) and that in Chichewa was 0.741 (95%CI: 0.604, 0.859). The cut-off with the highest Youden index was 1/2 in both Chitumbuka (sensitivity 45.5%, specificity of 83.4%, Youden Index 0.29) and Chichewa (sensitivity 62.5%, specificity 76.9%, Youden Index 0.39; Supporting Information **S4D table**).

**Figure 3:**
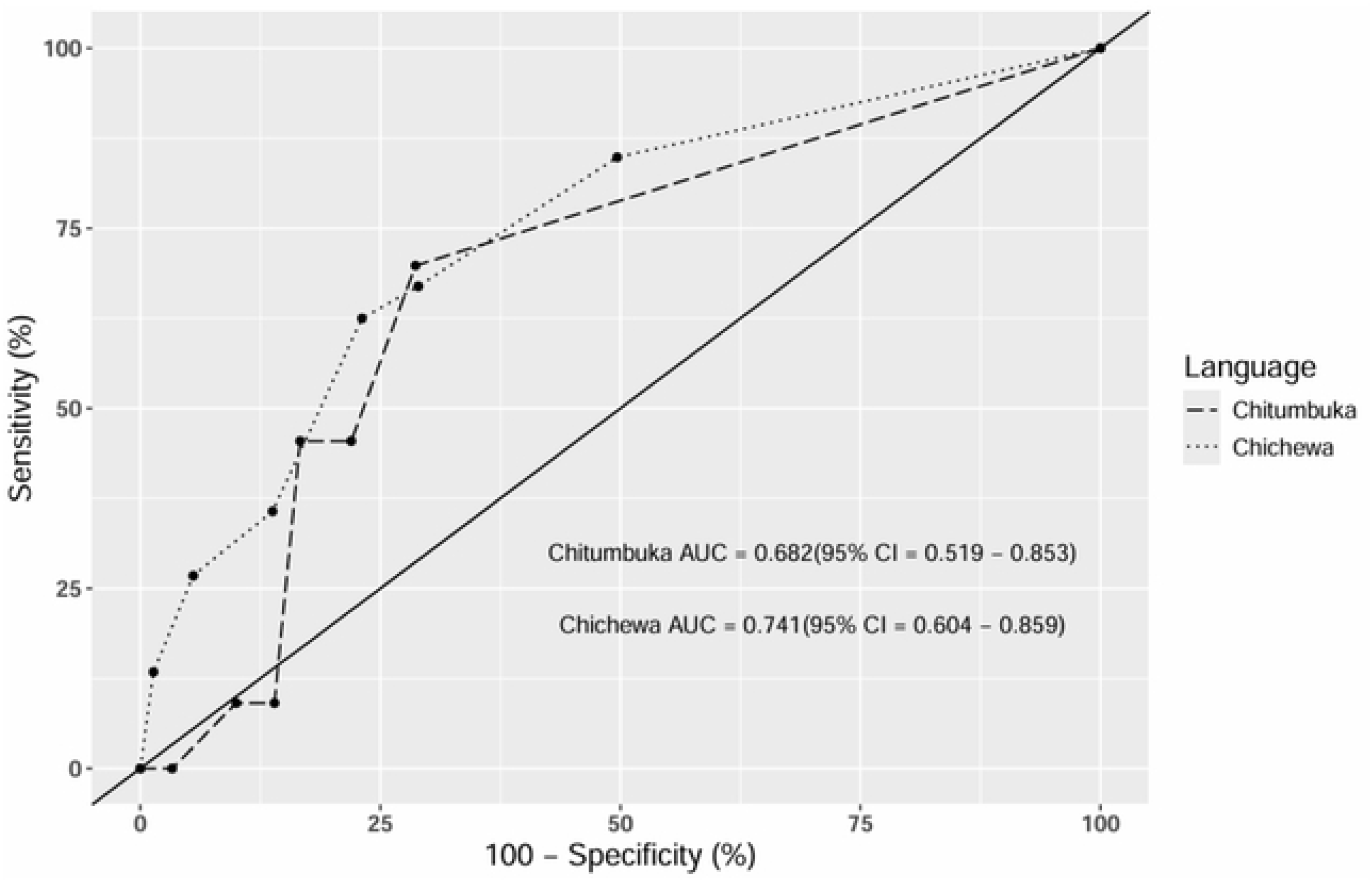
ROC curves for PC-PTSD-5 vs Post Traumatic Stress Disorder reference diagnosis in Chitumbuka and Chichewa.

## DISCUSSION

In this study we set out to translate, adapt and validate measures of anxiety, depression and PTSD symptoms for use in population-based epidemiological studies in Malawi. We developed Chitumbuka and Chichewa versions of the GAD-7, PHQ-9 and PC-PTSD-5 that were feasible to administer, and that demonstrated face and content validity (although less so for the PC-PTSD-5), alongside internal consistency and convergent validity. In both languages, GAD-7 and PC-PTSD-5 best fit a single factor structure, whilst the optimal model for PHQ-9 was a 2-factor structure. We found only partial measurement invariance between the 2 language versions of each measure. Criterion validation against DSM diagnoses showed AUCs for GAD-7 and PHQ-9 that were acceptable or good, except for the Chitumbuka version of the PHQ-9 for detection of MDE which was poor. AUC for PC-PTSD-5 was borderline acceptable in both languages.

### Strengths and Limitations

A strength of the study was the careful process of translation and adaptation conducted before the final version was adopted, with consultation with topic experts, field staff and community participants. A limitation was our change of PTSD measure from PCL-5 to the PC-PTSD-5 which meant that community participant FGDs and cognitive interviewing were not done specifically for the PC-PTSD-5. However, translations were discussed with experienced fieldworkers and the measure was piloted before use. Both fieldworkers and SCID interviewers were trained and supervised to promote reliability, however, formal inter-rater reliability testing of the was not done.

To ensure representation of a broad range of population groups (consistent with the Healthy Lives Malawi and Generation Malawi cohort studies), we purposively sampled men, antenatal women and postnatal women, and within each subgroup we recruited convenience samples. We acknowledge that the overall sample may not necessarily be representative of the population as a whole, thus limiting generalisability especially of the prevalence estimates. Our sample size of 800 screened/470 diagnostic interview was toward the higher end for this type of study [9]. However, confidence intervals around the AUCs are wide, and certainty in identifying the optimum cut-off scores is not high. We note that a large number of participants invited for SCID refused or could not be located, especially in the urban site, although this had minimal effect on the modelling.

### GAD-7

The GAD-7 demonstrated consistently good test characteristics, with robust face and content validity and only limited changes were made in administration of the measure, maximising technical equivalence with the original. Our data were consistent with previous GAD-7 validation studies in SSA finding acceptable to good internal consistency and evidence of convergent validity. We identified a single factor structure in both languages, consistent with most previous studies [11, 12, 15, 18, 19] and not the previously reported 2-factor structure [14, 17]. Finally, the GAD-7 demonstrated acceptable/good criterion validity for identification of Generalised Anxiety Disorder, again consistent with previous SSA GAD-7 criterion validation studies [13, 16, 20, 21].

### PHQ-9

The PHQ-9 had face/content validity in both languages. A significant technical change was the splitting of the psychomotor retardation/agitation question. We maintained the total score maximum of 27 by only counting the highest of the 2 item scores, but we note that this complicates the scoring, potentially limiting the use of this version to research contexts. For construct validity, both languages demonstrated acceptable internal consistency and evidence of convergent validity, similar to most previous validations in SSA. The factor structure of the PHQ-9, however, differed from previous studies which have generally identified a unidimensional structure [12, 15, 25, 29, 31, 33, 38, 39, 41, 42, 44–51, 53, 55], rather than a 2 factor [41], 3 factor [34], or variable 1 or 2 factor [14] structure. Consistent with Appiah et al 2020 [41], we identified a 2-factor structure in both languages, although the items loading onto each factor differed between the Chichewa and Chitumbuka. In Chichewa, one of the factors consisted of biological symptoms of depression (sleep, energy, appetite), whilst, in Chitumbuka, one of the factors could be understood as a severity indicator (suicidality, psychomotor retardation/agitation). The difference between the 2 language versions may be explained by contextual sources of variance in the expression of depression and mental health distress.

The PHQ-9 in both languages was accurate in detecting any depression (major or minor) and also for major depression in Chichewa. In previous criterion validation studies in SSA, 19 of 21 studies had AUC>0.8. Only a study in Uganda [34], and an underpowered study in South Africa [53] had AUC below this. Our findings are consistent with previous PHQ-9 validation in Chichewa-speaking attendees at an NCD clinic [40]. Our finding of poor accuracy for detection of MDE in Chitumbuka was surprising. It is possible that there was some misclassification in the SCID diagnosis of people on the borderline between minor and major depression in the Chitumbuka-speaking site. Also, it may reflect generally lower accuracy of Chitumbuka translated measures, perhaps reflecting contextual factors in a predominantly rural area. Previous studies have found that mental health screening tools tend to perform better in urban settings, at least partially attributable to higher urban literacy levels and greater population level exposure to “WEIRD” (Western Educated Industrialised Rich Democratic) societal concepts of mental health in urban settings, compared with rural locations. We did not find that the addition of 3 somatic complaint items (headache, general body pains and gastrointestinal symptoms) to the PHQ-9 improved the sensitivity/specificity for detection of DSM depressive episode. This is a similar finding to Habtamu et al. (2022) in an Ethiopian study who found that cognitive and emotional items in screening tools (PHQ-9, PHQ-15) were more strongly associated with depression diagnosis than physical symptoms [49].

### PC-PTSD-5

The PC-PTSD-5 proved more difficult to adapt to the Malawian setting and performed only moderately well in criterion validation in both languages/settings. We have described some missed steps in the adaptation process but these are unlikely to fully explain the performance limitations of the measure. Regarding face/content validity, it was difficult to be confident that respondents were reporting only *life threatening* traumatic events. We also noted that, in Karonga, 54% reported that a relative had died by suicide or other violent means suggesting some misunderstanding of the prompt. To improve recall accuracy, in a study in Mozambique, Massinga et al [58] changed the time period to ask about traumatic event in the previous month only, however, we decided that this would refocus the measure onto acute stress rather than more longstanding PTSD symptoms. Therefore, we maintained the opening question about experience of *lifetime* trauma (adding only the traumatic childbirth description given the intended use of the measure in the Generation Malawi birth cohort). Construct validity findings reported good internal consistency, a single factor structure and moderate convergence with the GAD-7 and PHQ-9 scores, similar to previous SSA validations [15, 16, 57, 58]. Our study is also the first SSA study to report AUC for the PC-PTSD-5. Both Chitumbuka and Chichewa versions showed similar low/moderate accuracy. It has been noted that DSM PTSD may be a more culture bound disorder than depression and anxiety, given the genesis of the construct in the particular circumstances of US Vietnam war veterans [76]. However, validation of longer PTSD measures in SSA settings have shown better test characteristics [77, 78], indicating that they may be a particular problem with ultra-brief PC-PTSD-5 measure that condenses all the syndrome symptoms into 5 compound yes/no items.

### Measurement invariance

Tests of measurement invariance indicated that configural invariance was supported for all three measures. Regarding metric invariance, for the PHQ-9, despite the marginal change in CFI, other fit statistics (RMSEA, TLI, and SRMR) supported invariance, so overall, there was evidence supporting metric invariance. For the GAD-7, the change in CFI exceeded the recommended threshold, and other fit statistics were all on the borderline of the recommended threshold, therefore, not supporting invariance. For PC-PTSD, the change in CFI also exceeded the recommended threshold; although other fit statistics were within the recommended thresholds, there was not enough evidence to support full metric invariance. Scalar invariance was not supported for any of the measures. Altogether, these findings indicate that comparisons of means across language groups should be interpreted with caution. Few studies have tested measurement invariance of the mental health screening measures that are administered in different languages within the same study. In a study similar to ours that validated English and Ixhosa versions of the same 3 measures amongst Adolescents in South Africa, Rodriquez et al [15] found that the GAD-7 and PHQ-9 demonstrated configural, metric, and scalar invariance across English and Ixhosa. For the PC-PTSD, the factor structure was comparable across languages for the configural model, the metric model was not assessed and the scalar invariance was not achieved.

### Choice of cut-off scores

Choice of optimal cut-off score depends on how the measure is being used (i.e. research vs clinical) and the balance of sensitivity to specificity required. When reporting prevalence figures for mental health conditions using screening tools, it is critical to report the test characteristics of the measures at the chosen cut-off. Using the Youden index, we identified cut-offs consistent across both language versions: GAD-7 cut-off 5/6, PHQ-9 cut-off 8/9, and PC-PTSD-5 cut-off 1/2. However, the balance of sensitivity and specificity at these cut-offs differs between the 2 languages; taking into account our finding of only partial measurement invariance between the 2 languages, this indicates some caution is needed in direct comparisons.

### Clinical utility of the measures

This study was conducted to develop and evaluate mental health measures for use in large population-based cohort studies, rather than for use as clinical screening tools. Nevertheless, the good test characteristics of the GAD-7 indicate that it might be a useful screening tool for Generalised Anxiety Disorder in Malawi where there are currently no locally validated clinical tools. Clinical use in this context should be as a screener leading to a diagnostic assessment. Regarding our version of the PHQ-9, the splitting of item 8 (psychomotor retardation/agitation) may make it prone to miscalculation of the total score in routine clinical use but is suitable for use in research contexts. Given the relatively poor performance of the PC-PTSD-5, it is difficult to recommend its use in clinical care at this time.

## Data Availability

The data are not made publicly available because they contain potentially identifiable clinical and demographic information that could compromise participant confidentiality. Furthermore, public data sharing via a data repository was not included in the approved ethics protocol for this study. De-identified data may be made available to qualified researchers upon reasonable request and following approval by the MEIRU data access committee. Please for data access enquiry.

